# Diagnostic accuracy study of the multiplex Truenat MTB Ultima/COVID-19 assay for simultaneous detection of Tuberculosis and SARS-CoV2 (COVID-19)

**DOI:** 10.64898/2026.01.08.26343747

**Authors:** Hafsah Tootla, Rita Székely, James Sserubiri, Widaad Zemanay, Mala Patidar, Moses Joloba, Cesar Ugarte-Gil, Morten Ruhwald, Manju Purohit, Helen Cox, Carlos Zamudio, Willy Ssengooba, Adam Penn-Nicholson, the COMBO study team

## Abstract

The COVID-19 pandemic led to significantly disrupted tuberculosis case detection and management. Additionally, overlapping symptoms, radiological findings and risk factors make differentiating tuberculosis and COVID-19 disease difficult.

We conducted a prospective multicentre diagnostic accuracy study to determine sensitivity, specificity and operational characteristics of the Truenat MTB Ultima/COVID-19 assay, using a combined sputum plus nasopharyngeal swab specimen, in a single multiplex molecular assay. Participants were consenting adults with presumptive tuberculosis enrolled via convenience sampling from Uganda, Peru, South Africa, and India between August 2022 and December 2023. A microbiological reference standard of sputum GeneXpert MTB/RIF Ultra and culture was used for tuberculosis, whereas national RT-PCR was used for COVID-19. Wilson’s score method was used to determine the sensitivity and specificity.

Of 1,928 participants enrolled, median age was 38 years (IQR 28-50), 359/1928 (18.6%) previously had TB, and 287/1928 (14.9%) were HIV-positive. Overall prevalence of tuberculosis was 24.8% [95% CI 22.9-26.8%]. Prevalence of COVID-19 was 3.8% [3.1-4.8%] overall, and 4.7% [3.1-7.0%] in those with confirmed tuberculosis. Overall sensitivity of Truenat MTB Ultima/COVID-19 for tuberculosis was 79.8% [95% CI 76.0-83.2%]. When comparing paired samples, Truenat MTB Ultima/COVID-19 had a 9.5% [6.5-13.1%] decreased sensitivity against sputum TB culture, compared to GeneXpert Ultra; (82.8% [ 78.9-86.1%] vs 92.4% [89.4-94.5%]). Overall specificity of Truenat MTB Ultima/COVID-19 for tuberculosis was 98.9% [98.2-99.3%]. For COVID-19 detection, sensitivity of Truenat MTB Ultima/COVID-19 was 64.4% [52.9-74.4%], with specificity of 99.2% [98.7-99.5%].

Although optimal diagnostic performance was not demonstrated, the potential and need for rapid development of tests that integrate tuberculosis diagnosis with detection of other relevant respiratory infections is highlighted.

The study was registered on ClinicalTrials.gov (NCT05405296).

## INTRODUCTION

For many decades, tuberculosis (TB) has been the leading cause of infectious death, temporarily surpassed by the SARS-CoV2 (COVID-19) virus during the COVID-19 pandemic, and now most recently re-emerging once again as the number one cause of death by infectious disease [1,2].

Intensified research and innovation in TB control, with a focus to meet the WHO End TB strategy targets (by the year 2030) of 90% reduction in TB deaths, 80% reduction in TB incidence rate, and no TB-affected families facing catastrophic costs due to TB, have called for integrated person-centred TB care and have resulted in the development of revolutionary rapid molecular TB diagnostic tests, and shorter effective oral treatment and preventative strategies [3]. Despite these powerful advancements, major gaps, particularly in TB case detection (including affordability and accessibility) and treatment coverage, still exist [1,3]. Over 10 million people per year still develop TB disease and ending the TB epidemic remains an urgent priority [1,3,4]. Although substantial progress had been made with TB control in the last decade, the COVID-19 pandemic caused significant setbacks [1,4–6]. TB case detection dropped by ∼20%, access to TB services during the pandemic was limited and there was an overall ∼20% reduction in the number of people receiving TB preventative therapy [5–7]. Tuberculosis prevalence amongst hospitalised COVID-19 cohorts was estimated at between 2-8% in high TB-burden settings and overlapping clinical symptoms, radiological features and onset of symptoms, particularly in TB-endemic countries, made differentiating TB and COVID-19 disease difficult [7–10]. Furthermore, shared risk factors for TB and COVID-19 infection and disease, such as overcrowded living spaces and comorbidities like HIV and diabetes, as well as co-infection with both TB and COVID-19, added complications in making an accurate diagnosis of TB when resources were being redirected towards diagnosis and containment of COVID-19 [5].

The COVID-19 pandemic brutally demonstrated the consequences of disrupted TB diagnosis and care, and highlighted the need for rapid integrated testing strategies to diagnose and limit TB transmission, even when new threats like the COVID-19 pandemic occur.

The development of the Truenat^TM^ MTB Ultima/COVID-19 multiplex PCR was in response to the unmet need for a single test capable of diagnosing both important high morbidity and high mortality infectious diseases. We conducted a multi-country study to determine the diagnostic accuracy of the Truenat MTB Ultima/COVID-19 assay for the detection of both TB and COVID-19 against a TB and COVID-19 microbiological reference standard (MRS), respectively, and against a TB comparator assay currently in use in each country.

## MATERIALS AND METHODS

### Study design and population

This prospective multinational diagnostic accuracy study enrolled adults (age>18 years) by convenience sampling from participating healthcare facilities in Uganda (Makerere University, Kampala) between 31 August 2022 and 05 December 2023; in Peru (Instituto de Medicina Tropical Alexander von Humboldt, Universidad Peruana Cayetano Heredia, Lima) between 26 September 2022 and 05 December 2023; South Africa (Site B Clinic, Khayelitsha, Cape Town) between 19 October 2022 and 05 December 2023; and in India (RD Gardi Medical College, Ujjain, Madhya Pradesh) between 20 December 2022 and 05 December 2023. Once all sites had achieved their sample size target, participants were enrolled using competitive enrolment.

Participants were approached for enrolment if they reported ≥ 1 of the following symptoms suggestive of TB: cough ≥ 2 weeks, fever, night sweats or unintended weight-loss. Participants were excluded from the study if they received any prior TB treatment within 60 days of enrolment or any prior TB preventative therapy within 6 months of enrolment, were unable to provide the required first-day samples, or if all the required samples had not been collected before a 3rd dose of TB treatment was taken. Samples with indeterminate or missing test results (either index or reference) were excluded from the primary analysis.

Participants provided written informed consent, and the study received ethical approval from all participating sites (Uganda: Makerere University School of Biomedical Sciences Research Ethics Committee (MAKSBS-REC: SBS-2022-122) and the Uganda National Council for Science and Technology (UNCST#HS2346ES); Peru: Human Research Ethics Committee of the Universidad Peruana Cayetano Heredia (SIDISI 207221); India: RD Gardi Medical College IEC (IEC 05-06/2022); South Africa: Human Research Ethics Committee of the University of Cape Town (HREC 070/ 2022). The study was registered with ClinicalTrials.gov (NCT05405296), and in Peru the study was registered with the Peruvian National Institute of Health Repository PRISA (number 2573). The study protocol has been previously published [11].

### Primary and Secondary outcomes

The primary outcome for this study was to determine the diagnostic accuracy of Truenat MTB Ultima/COVID-19 for TB detection compared to a defined TB microbiological reference standard (MRS).

Secondary outcomes were 1) to determine the diagnostic accuracy of Truenat MTB Ultima/COVID-19 for COVID-19 detection compared to a COVID-19 MRS, 2) to determine the diagnostic accuracy of Truenat MTB Ultima/COVID-19 for TB detection compared to GeneXpert Ultra using TB culture as the reference standard, 3) to determine the prevalence of COVID-19 amongst all study participants, 4) to determine the prevalence of COVID-19 amongst participants with confirmed TB based on a positive TB MRS, 5) to determine the diagnostic accuracy of Truenat MTB Ultima/COVID-19 for TB detection and COVID-19 detection using an alternative sample of tongue and mid-turbinate swab compared to the TB and COVID-19 MRS respectively.

### Sample collection and Testing

On Day 1 of enrolment, a healthcare worker collected two nasopharyngeal swabs for COVID-19 testing with the COVID-19 MRS and Truenat MTB Ultima/COVID-19 multiplex PCR respectively. For TB testing, a minimum of 3 ml of homogenised sputum, split for testing with the Truenat MTB Ultima/COVID-19 PCR (1 ml) and TB MRS (2 ml), respectively, was collected. One healthcare-worker-collected mid-turbinate swab and one healthcare-worker-collected tongue swab (the alternative and easier-to-collect sample types compared to nasopharyngeal swab and sputum) was collected for both COVID-19 and TB testing. Three additional healthcare-worker-collected tongue swabs were collected for biobanking. Baseline demographic and clinical data were captured, and diabetes and hypertension screening performed. Participants were sent home with a labelled empty sputum sample container for next day sample collection.

On Day 2 (or within 7 days of enrolment provided that all the required samples were collected before a 3^rd^ dose of TB treatment was taken), participants collected 2 ml of early morning sputum at home, and brought this to the facility for TB testing. At the facility, participants self-swabbed two tongue swabs while being observed by a healthcare worker. Thereafter two additional healthcare-worker-collected tongue swabs were also collected. All tongue swabs from Day 2 were stored for biobanking. Biobanked samples, and where available, digital chest X-rays that were anonymised after clinician review, were collected for storage in a databank for future research and analysis.

Clinical information and reference test results were not available to laboratory staff performing the index test. Clinical information and index test results were not available to laboratory staff performing the reference tests. Index test results from this research study were not available to clinicians or used for clinical care.

### Truenat MTB Ultima/COVID-19 multiplex PCR assay description

The assay was performed according to manufacturer instructions. Briefly, 500 µl of liquified expectorated sputum was added directly to a vial containing a nasopharyngeal swab in 500 µl Molbio Viral Transport Medium (VTM), mixed, and 500 µl transferred to the Truenat sample pretreatment buffer. Nucleic acids were extracted from the total volume using Trueprep® AUTO/AUTO v2 Sample Prep Device and Trueprep AUTO/AUTO v2 Universal sample prep kit. For the alternative sample secondary outcome, a tongue swab was added directly to a tube containing a mid-turbinate nasal swab in 500 µl VTM, and the full volume was similarly extracted on the Trueprep platform. Thereafter, 6 µl of extracted nucleic acid was dispensed into a microtube containing the freeze-dried PCR reagents. After allowing ∼30-60 seconds for the dried PCR reagents to rehydrate with the nucleic acids from the sample, 6 μL of clear suspension was pipetted and dispensed into the reaction well of the Truenat MTB Ultima/COVID-19 chip. The chip was then inserted in the Truelab Real Time micro–PCR Analyzer for simultaneous TB and COVID-19 detection.

### Microbiological reference tests

The TB microbiological reference standard (MRS) for the primary outcome of TB detection from sputum (with Truenat MTB Ultima/COVID-19) was a composite of sputum GeneXpert MTB/RIF Ultra (first day), and sputum MGIT and LJ culture (first and second day).

The TB MRS for the secondary outcome of TB detection from sputum (with Truenat MTB Ultima/COVID-19) when compared to TB detection from sputum by the GeneXpert Ultra was sputum MGIT and LJ culture (first and second day).

The COVID-19 reference standard for the secondary outcome of COVID-19 detection from nasopharyngeal swab (with Truenat MTB Ultima/COVID-19) was a country approved RT-PCR (first day).

### Data and statistical analysis

The sample size target was determined based on the study enrolling an overall minimum of 270 participants with confirmed TB. Using an estimated TB prevalence of 20%, with ∼10% of patients assumed to be lost to follow up, the total sample size required for the study was 1,480, with a minimum sample size target of 370 participants for each site.

Descriptive statistics on patient characteristics and estimates of diagnostic accuracy stratified by site, sputum smear status, previous TB history and previous HIV status diagnosis was performed. Categorical variables are summarised as absolute numbers and relative (percent) frequencies. Continuous variables are summarised as medians with interquartile ranges (25th and 75th percentiles). Wilson’s score method was used to determine the sensitivity and specificity and presented as point estimates with two-sided 95% confidence intervals. The analysis was performed using SAS® System (Version 9.4 or higher) or R statistical language (version 3.4.0 or higher) and Microsoft Excel 2017 (version 15.34 or higher).

## RESULTS

Between 31 August 2022 and 05 December 2023, 1,984 participants were enrolled, with 1,928 included in the analysis after exclusion of 56 participants (Figure 1). The median age of participants was 38 years (IQR 28-50 years), 47.6% (n/N = 918/1928) were female,18.6% (359/1928) had previous TB, and 14.9% (287/1928) reported or were diagnosed with HIV. The overall prevalence of diabetes and hypertension was 103/1926 (5.3%, 95% Confidence Interval 4.4-6.4%) and 114/1924 (5.9%, CI 5-7.1%) respectively. The overall prevalence of TB using the primary MRS for TB detection was 478/1928 (24.8%, CI 22.9-26.7%). Samples from all participants underwent sputum smear microscopy; of those who were microbiologically confirmed with TB by the primary MRS, 335/478 (70.1%) were sputum smear positive, ranging from 54/86 (62.8%) in Uganda to 125/153 (81.7%) in Peru. A total of 1588/1928 (82.4%) of all participants who underwent sputum smear microscopy were smear negative. During the study period, each country had a differing prevalence of COVID-19 based on country-specific waves. The overall prevalence of COVID-19 was 73/1910 (3.8%, CI 3.1-4.8%), with the prevalence of COVID-19 in those with confirmed TB at 22/470 (4.7%, CI 3.1-7%) (Table 1).

**Figure 1:**
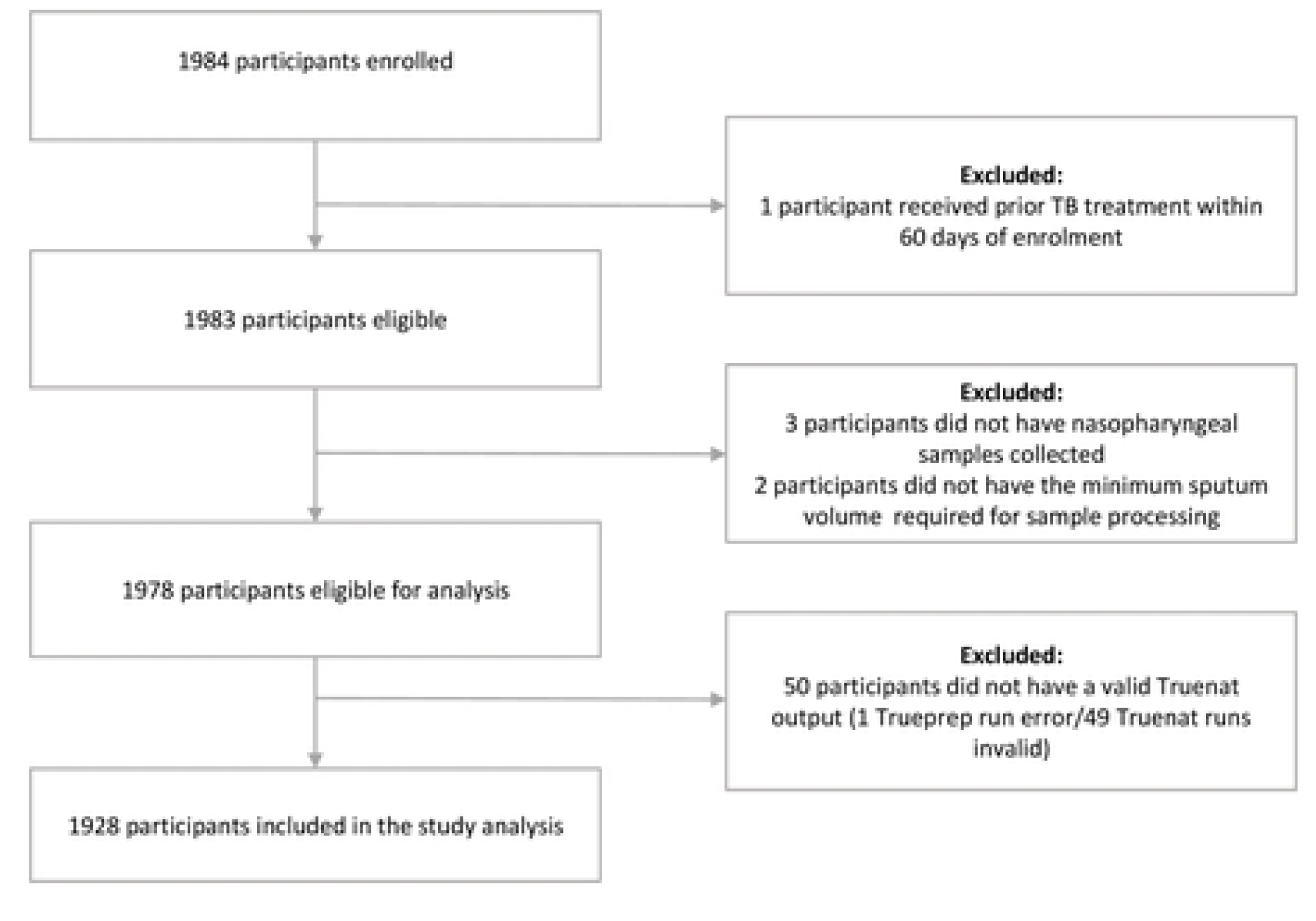
STARO diagram representationof panicipants enrolled, panicipants excluded, and participants included in thestudy analyses

**Table 1:** Demographic, clinical and laboratory characteristics of participants in the study analysis.

|  | Total<br>N=1928 (%) | Uganda<br>N=611 (31.7%) | Peru<br>N=504 (26.1%) | South Africa<br>N=426 (22.1%) | India<br>N=387 (20.1%) |
| --- | --- | --- | --- | --- | --- |
| **Gender: n (%)** |  |  |  |  |  |
| Male | 1008 (52.3%) | 295 (48.3%) | 245 (48.6%) | 238 (55.9%) | 230 (59.4%) |
| Female | 918 (47.6%) | 316 (51.7%) | 257 (51.0%) | 188 (44.1%) | 157 (40.6%) |
| Undifferentiated | 2 (0.1%) | - | 2 (0.4%) | - | - |
| **Age in years (IQR)** |  |  |  |  |  |
|  | 38 (28.0-50.0) | 32 (24.0-43.0) | 41 (27.0-58.0) | 38 (31.0-47.0) | 46 (30.0-60.0) |
| **Previous history of TB n (%)** |  |  |  |  |  |
| No | 1559 (80.9%) | 552 (90.3%) | 383 (76.0%) | 279 (65.5%) | 345 (89.1%) |
| Yes | 359 (18.6%) | 55 (9.0%) | 121 (24.0%) | 147 (34.5%) | 36 (9.3%) |
| Did not know | 10 (0.5%) | 4 (0.7%) | 0 (0%) | 0 (0%) | 6 (1.6%) |
| **Previous diagnosis of HIV n (%)** |  |  |  |  |  |
| No | 1521 (78.9%) | 455 (74.5%) | 403 (80.0%) | 278 (65.3%) | 385 (99.5%) |
| Yes | 287 (14.9%) | 126 (20.6%) | 14 (2.8%) | 146 (34.3%) | 1 (0.3%) |
| Did not know | 120 (6.2%) | 30 (4.9%) | 87 (17.3%) | 2 (0.5%) | 1 (0.3%) |
| **Overall TB prevalence (N=1920)** |  |  |  |  |  |
| n/N (%)<br>(95%CI) | 476/1920 (24.8%)<br>(22.9-26.8%) | 86/606 (14.2%)<br>(11.6-17.2%) | 151/501 (30.1%)<br>(26.3-34.3%) | 130/426 (30.5%)<br>(26.3-35.0%) | 109/387 (28.2%)<br>(23.9-32.8%) |
| **Microscopy Smear status for TB MRS positive participants (N=478) n/N (%)** |  |  |  |  |  |
| MRS-positive:<br>Smear negative | 143/478 (29.9%) | 32/86 (37.2%) | 28/153 (18.3) | 48/130 (36.9%) | 35/019 (32.1) |
| MRS-positive:<br>Smear positive | 335/478 (70.1%) | 54/86 (62.8%) | 125/153 (81.7%) | 82/130 (63.1%) | 74/109 (67.9%) |
| Scanty | 58/335 (17.3%) | 0/54 (0.0%) | 40/125 (32.0%) | 5/82 (6.1%) | 13/74 (17.6%) |
| 1+ | 111/335 (33.1%) | 10/54 (18.5%) | 31/125 (24.8%) | 27/82 (32.9%) | 43/74 (58.1%) |
| 2+ | 78/335 (23.3%) | 22/54 (40.7%) | 25/125 (20.0%) | 15/82 (18.3%) | 16/74 (21.6%) |
| 3+ | 88/335 (26.3%) | 22/54 (40.7%) | 29/125 (23.2%) | 35/82 (42.7%) | 2/74 (2.7%) |
| **Overall COVID-19 prevalence (N=1910)** |  |  |  |  |  |
| n/N (%)<br>(95%CI) | 73/1910 (3.8%)<br>(3.1%-4.8%) | 4/611 (0.7%)<br>(0.3%-1.7%) | 29/497 (5.8%)<br>(4.1-8.3%) | 21/415 (5.1%)<br>(3.3-7.6%) | 19/387 (4.9%)<br>(3.2-7.5%) |
| **COVID-19 prevalence in TB MRS positive participants (N=470)** |  |  |  |  |  |
| n/N (%)<br>(95%CI) | 22/470 (4.7%)<br>(3.1-7.0%) | 1/86 (1.2%)<br>(0.2-6.3%) | 10/149 (6.7%)<br>(3.7-11.9%) | 6/126 (4.8%)<br>(2.2-10.0%) | 5/109 (4.6%)<br>(2.0-10.3%) |
| **Overall Diabetes prevalence (N=1926)** |  |  |  |  |  |
| n/N (%)<br>(95%CI) | 103/1926 (5.3%)<br>(4.4-6.4%) | 15/611 (2.5%)<br>(1.5-4.0%) | 50/504 (9.9%)<br>(7.6-12.8%) | 28/424 (6.6%)<br>(4.6-9.4%) | 10/387 (2.6%)<br>(1.4-4.7%) |
| **Overall Hypertensive prevalence (N=1924)** |  |  |  |  |  |
| n/N (%)<br>(95%CI) | 114/1924 (5.9%)<br>(5.0-7.1%) | 50/607 (8.2%)<br>(6.3-10.7%) | 19/504 (3.8%)<br>(2.4-5.8%) | 42/426 (9.9%)<br>(7.4-13.1%) | 3/387 (0.8%)<br>(0.3%-2.3%) |

N: Number of evaluated participants. n/N: Number of variable-positive participants/ number of evaluated participants. IQR: interquartile range; 95% CI: 95% Confidence Interval; TB MRS: TB primary microbiological reference standard of a composite of sputum GeneXpert MTB/RIF Ultra (first day), and sputum MGIT and LJ culture (first and second day)

During the diagnostic accuracy study, no Severe Adverse Events (SAEs) were reported.

### Diagnostic performance for TB diagnosis

The overall sensitivity of the Truenat MTB Ultima/COVID-19 assay for detection of TB from sputum was 380/476 (79.8%, CI 76.0-83.2%) when compared to the TB MRS. Sensitivity was better in sputum smear-positive participants at 310/333 (93.1%, CI 89.8-95.4%) (Table 2), and in India at 101/109, (92.7%, CI 86.2-96.2%) when compared to the TB MRS (Table 3). Overall specificity of the Truenat MTB Ultima/COVID-19 when compared to the TB MRS was 1428/1444 (98.9%, CI 98.2-99.3%) (Table 2).

**Table 2:** Diagnostic performance of Truenat MTB Ultima/COVID-19 for TB and COVID-19 detection among adults with TB symptoms compared to a composite reference of sputum GeneXpert Ultra, MGIT and LJ culture for TB, and a national site-specific RT-PCR for COVID-19, using (a) sputum and nasopharyngeal swab and (b) tongue and mid-turbinate swab.

|  | N | TP | FP | FN | TN | Sensitivity %<br>(95% CI) | * Sensitivity %<br>smear-positive<br>(95% CI) (n/N) | *Sensitivity %<br>smear-negative<br>(95% CI) (n/N) | Specificity %<br>(95% CI) |
| --- | --- | --- | --- | --- | --- | --- | --- | --- | --- |
| (a) Sputum + nasopharyngeal swab |  |  |  |  |  |  |  |  |  |
| MTB | 1920 | 380 | 16 | 96 | 1428 | 79.8%<br>(76.0-83.2) | 93.1%<br>(89.8-95.4)<br>(n= 310/333) | 49.0%<br>(40.9-57.1)<br>(n=70/143) | 98.9%<br>(98.2-99.3) |
| COVID-19 | 1902 | 47 | 15 | 26 | 1814 | 64.4%<br>(52.9-74.4) | NA | NA | 99.2%<br>(98.7-99.5%) |
| (b) Tongue swab + mid-turbinate swab |  |  |  |  |  |  |  |  |  |
| MTB | 837 | 94 | 7 | 111 | 625 | 45.9%<br>(39.2-52.7) | 63.7%<br>(55.3-71.3%<br>(n=86/135) | 10.9%<br>(5.4-20.9)<br>(n=7/64) | 98.9%<br>(97.7-99.5) |
| COVID-19 | 832 | 5 | 2 | 7 | 818 | 41.7%<br>(19.3-68.0) | NA | NA | 99.8%<br>(99.1-99.9) |

N: Number of evaluated participant samples; TP: True Positive; FP: False Positive; FN: False Negative; TN: True Negative; 95% CI: 95% Confidence Interval; n/N: Number of test-positive participants / Number of evaluated participants; NA: not applicable
\*Smear-status of TB primary microbiological reference standard-positive participants. (TB primary microbiological reference standard is a composite of sputum Gene Xpert MTB/RIF Ultra (first day), and sputum MGIT and LJ culture (first and second day))

**Table 3:** Diagnostic performance of Truenat™ MTB Ultima/COVID-19for TB detection among adults with TB symptoms compared to a composite reference of sputum GeneXpert Ultra, MGIT and LJ culture using (a) sputum and nasopharyngeal swab and (b) tongue and mid-turbinate swabs stratified by previous HIV diagnosis, previous TB history and country.

|  | Number of evaluated participant samples | Sensitivity % (95% CI) (n/N) | Specificity % (95% CI) (n/N) |
| --- | --- | --- | --- |
| **(a) Sputum and nasopharyngeal swab** |  |  |  |
| **Previous diagnosis of HIV** |  |  |  |
| Yes | 286 | 73.9% (62.5-82.8) (n=51/69) | 98.6% (96.0-99.5) (n=214/217) |
| No | 1515 | 81.3% (77.0-84.9) (n=304/374) | 99.0% (98.3-99.5) (n=1130/1141) |
| **Previous history of TB** |  |  |  |
| Yes | 357 | 78.0% (69.3-84.7) (n=85/109) | 98.4% (95.9-99.4) (n=244/248) |
| No | 1553 | 80.3% (75.9-84.1) (n=294/366) | 99.0% (98.2-99.4) (n=1175/1187) |
| **Country** |  |  |  |
| Uganda | 606 | 82.6% (73.2-89.1) (n=71/86) | 99.4% (98.3-99.8) (n=517/520) |
| Peru | 501 | 73.5% (66.0-79.9) (n=111/151) | 99.4% (97.9-99.8) (n=348/350) |
| South Africa | 426 | 74.6% (66.5-81.3) (n=97/130) | 98.6% (96.6-99.5) (n=292/296) |
| India | 387 | 92.7% (86.2-96.2) (n=101/109) | 97.5% (94.9-98.8) (n=271/278) |
| **(b) Tongue swab and mid-turbinate nasal swab** |  |  |  |
| **Previous diagnosis of HIV** |  |  |  |
| Yes | 161 | 42.4% (27.2-59.2) (n=14/33) | 100% (97.1-100) (n=128/128) |
| No | 612 | 49.0% (41.3-56.8) (n=77/157) | 98.7% (97.2-99.4) (n=449/455) |
| **Previous history of TB** |  |  |  |
| Yes | 170 | 44.4% (30.9-58.8) (n=20/45) | 99.2% (95.6-99.9) (n=124/125) |
| No | 665 | 46.3% (38.7-54.0) (n=74/160) | 98.8% (97.4-99.5) (n=499/505) |
| **Country** |  |  |  |
| Uganda | 356 | 51.6% (39.4-63.6) (n=32/62) | 99.0% (97.0-99.7) (n=291/294) |
| Peru | 225 | 33.8% (23.9-45.4) (n=24/71) | 98.7% (95.4-99.6) (n=152/154) |
| South Africa | 218 | 52.2% (40.5-63.7) (n=35/67) | 100% (97.5-100) (n=151/151) |
| India | 38 | 60.0% (23.1-88.2) (n=3/5) | 93.9% (80.4-98.3) (n=31/33) |

95% CI: 95% Confidence Interval; Sensitivity n/N: Number of index test-positive participants / Number of composite reference test-positive participants; Specificity n/N: Number of index test-negative participants / Number of composite reference test-negative participants

When compared to GeneXpert Ultra (sensitivity 387/419 (92.4%, CI 89.4-94.5%)), the Truenat MTB Ultima/COVID-19 ((sensitivity 347/419 (82.8%, CI 78.9-86.1%)) had a 9.5% [CI 6.5-13.1%] decreased sensitivity for TB detection when compared against sputum TB culture. The largest reduction in performance was in Peru, with a reduction in sensitivity by 17.9% [CI 12.0%-25.2%], with the other sites showing a reduction in sensitivity by 1.2 to 9.6%. The decreased sensitivity in sputum smear negative participants was also high, with a reduction of 19.6% [CI 9.4-30.1%] in sensitivity (Table 4).

**Table 4:** Diagnostic accuracy of Truenat™ MTB Ultima/COVID-19 multiplex compared to GeneXpert Ultra for TB detection among adults with TB symptoms against a reference standard of sputum TB culture.

|  | Sensitivity<br>% (95% CI)<br>(n/N) | Specificity<br>% (95% CI)<br>(n/N) |
| --- | --- | --- |
| **Overall** | 1906 |  |
| Truenat MTB Ultima/Covid-19 | 82.8% (78.9% to 86.1%)<br>(n=347/419) | 96.7% (95.7% to 97.5%)<br>(n=1438/1487) |
| GeneXpert Ultra | 92.4% (89.4% to 94.5%)<br>(n=387/419) | 96.2% (95.1% to 97.1%)<br>(n=1431/1487) |
| Comparative: Truenat - Ultra | -9.5% (-13.1% to -6.5%) | 0.5% (-0.4% to 1.3%) |
| **Country** |  |  |
| **Uganda** | 605 |  |
| Truenat MTB Ultima/Covid-19 | 87.8% (78.5% to 93.5%)<br>(n=65/74) | 98.3% (96.8% to 99.1%)<br>(n=522/531) |
| GeneXpert Ultra | 90.5% (81.7% to 95.3%)<br>(n=67/74) | 97.7% (96.1% to 98.7%)<br>(n=519/531) |
| Comparative: Truenat - Ultra | -2.7% (-11.6% to 5.7%) | 0.6% (-0.7% to 1.9%) |
| **Peru** | 499 |  |
| Truenat MTB Ultima/Covid-19 | 75.2% (67.6% to 81.5%)<br>(n=109/145) | 98.9% (97.1% to 99.6%)<br>(n=350/354) |
| GeneXpert Ultra | 93.1% (87.8% to 96.2%)<br>(n=135/145) | 98.3% (96.4% to 99.2%)<br>(n=348/354) |
| Comparative: Truenat - Ultra | -17.9% (-25.2% to -12.0%) | 0.6% (-1.0% to 2.4%) |
| **South Africa** | 417 |  |
| Truenat MTB Ultima/Covid-19 | 80.0% (71.8% to 86.3%)<br>(n=92/115) | 97.0% (94.4% to 98.4%)<br>(n=293/302) |
| GeneXpert Ultra | 89.6% (82.6% to 93.9%)<br>(n=103/115) | 95.4% (92.4% to 97.2%)<br>(n=288/302) |
| Comparative: Truenat - Ultra | -9.6% (-16.3% to -5.4%) | 1.7% (-0.8% to 4.4%) |
| **India** | 385 |  |
| Truenat MTB Ultima/Covid-19 | 95.3% (88.5% to 98.2%)<br>(n=81/85) | 91.0% (87.2% to 93.7%)<br>(n=273/300) |
| GeneXpert Ultra | 96.5% (90.1% to 98.8%)<br>(n=82/85) | 92.0% (88.4% to 94.6%)<br>(n=276/300) |
| Comparative: Truenat - Ultra | -1.2% (-7.9% to 5.2%) | -1.0% (-3.6% to 1.4%) |
| **Smear Status of TB sputum culture-positive participants** |  |  |
| **Smear Positive** | 322 |  |
| Truenat MTB Ultima/Covid-19 | 93.2% (89.9% to 95.4%)<br>(n=300/322) | NA |
| GeneXpert Ultra | 99.7% (98.3% to 99.9%)<br>(n=321/322) | NA |
| Comparative: Truenat - Ultra | -6.5% (-9.9% to -4.0%) | NA |
| **Smear Negative** | 97 |  |
| Truenat MTB Ultima/Covid-19 | 48.5% (38.8% to 58.3%)<br>(n=47/97) | NA |
| GeneXpert Ultra | 68.0% (58.2% to 76.5%)<br>(n=66/97) | NA |
| Comparative: Truenat - Ultra | -19.6% (-30.1% to -9.4%) | NA |
| **Previous diagnosis of HIV** |  |  |
| **Previous positive HIV diagnosis** | 286 |  |
| Truenat MTB Ultima/Covid-19 | 78.7% (66.9% to 87.1%)<br>(n=48/61) | 97.3% (94.3% to 98.8%)<br>(n=219/225) |
| GeneXpert Ultra | 83.6% (72.4% to 90.8%)<br>(n=51/61) | 96.4% (93.1% to 98.2%)<br>(n=217/225) |
| Comparative: Truenat - Ultra | -4.9% (-14.4% to 3.1%) | 0.9% (-1.9% to 3.9%) |
| **Previous negative HIV diagnosis** | 1502 |  |
| Truenat MTB Ultima/Covid-19 | 84.0% (79.7% to 87.6%)<br>(n=274/326) | 96.5% (95.3% to 97.4%)<br>(n=1135/1176) |
| GeneXpert Ultra | 94.5% (91.4% to 96.5%)<br>(n=308/326) | 96.0% (94.7% to 97.0%)<br>(n=1129/1176) |
| Comparative: Truenat - Ultra | -10.4% (-14.6% to -6.8%) | 0.5% (CI -0.4 to 1.5%) |
| **Previous TB history** |  |  |
| **History of previous TB (yes)** | **354** |  |
| Truenat MTB Ultima/Covid-19 | 85.4% (76.6% to 91.3%)<br>(n=76/89) | 95.1% (91.8% to 97.1%)<br>(n=252/265) |
| GeneXpert Ultra | 92.1% (84.6% to 96.1%)<br>(n=82/89) | 92.5% (88.6% to 95.1%)<br>(n=245/265) |
| Comparative: Truenat - Ultra | -6.7% (-13.9% to -2.3%) | 2.6% (-0.3% to 6.0%) |
| **No history of prior TB** | **1542** |  |
| Truenat MTB Ultima/Covid-19 | 82.1% (77.6% to 85.8%)<br>(n=270/329) | 97.0% (95.9% to 97.8%)<br>(n=1177/1213) |
| GeneXpert Ultra | 92.4% (89.0% to 94.8%)<br>(n=304/329) | 97.0% (95.9% to 97.8%)<br>(n=1177/1213) |
| Comparative: Truenat - Ultra | -10.3% (-14.5% to -6.6%) | 0.0% (-0.8% to 0.8%) |

95% CI: 95% Confidence Interval; Sensitivity n/N: Number of test-positive participant samples / Number of reference test-positive participant samples; Specificity n/N: Number of test-negative participant samples / Number of reference test-negative participant samples; NA: Not applicable
Comparison of the Truenat MTB Ultima/COVID-19 assay to the GeneXpert Ultra assay to detect MTB was done against a secondary reference standard of sputum TB culture alone. Sample sizes here reflect the number of participants where samples provided valid results for all tests of Truenat MTB Ultima/COVID-19, GeneXpert Ultra and sputum TB culture.

The overall sensitivity of the Truenat MTB Ultima/COVID-19 assay for detection of TB from tongue and mid-turbinate nasal swabs was 94/205 (45.9%, CI 39.2-52.7%) compared to the TB MRS. Overall specificity was 625/632 (98.9%, CI 97.7-99.5%). Sensitivity was higher but still poor in smear-positive participants (86/135, 63.7%, CI 55.3-71.3) (Table 2).

### Diagnostic performance for COVID-19 diagnosis

The overall sensitivity of the Truenat MTB Ultima/COVID-19 assay for detection of COVID-19 from nasopharyngeal swabs when compared to a national site-specific RT-PCR was 47/73 (64.4%, CI 52.9-74.4%) (Table 2). Performance was better in India with a sensitivity of 18/19 (94.7%, CI 75.4-99.1%) (Supplementary Table 1). Overall Specificity was 1814/1829 (99.2%, CI 98.7-99.5%) (Table 2).

The overall sensitivity and specificity of the Truenat MTB Ultima/COVID-19 assay for detection of COVID-19 from tongue and mid-turbinate nasal swab was 5/12 (41.7%, CI 19.3-68.0%) and 818/820 (99.8%, CI 99.1-99.9%), respectively. (Table 2)

### Assay non-actionable results: error and invalid rates

Supplementary tables 2A and 2B details the number of non-actionable results for each specimen type. Of 1,980 Trueprep assays run using the sputum plus nasopharyngeal swab method, 19 (0.96%) resulted in an error or no result, and 2 had missing data. When repeated, 18/19 (95%) resolved. Extracted DNA from 1,977 participants was run on the Truenat assay, with 1,705 (86.2%) providing valid results. The 271 samples with invalid or no results were repeated, with 215 (79.3%) resolving on repeat testing. In total, 1,920/1,977 (97%) samples tested yielded actionable results.

When testing the alternative sample of tongue swab plus mid-turbinate nasal swab, 3/852 (0.35%) Trueprep run samples were no result/error, and 3 (0.35%) were not done. Of the 3 samples repeated, all resolved. When tested on Truenat, 104/849 (12.25%) were invalid, and 7 (0.82%) were no result/error. Upon repeat Truenat testing, 99/111 (89.19%) resolved. In total, 837/849 (98.6%) of samples tested yielded actionable results.

## DISCUSSION

The COVID-19 pandemic had a large negative impact on TB prevention and treatment [1,4–7]. Decades of progress made with TB control and elimination was set back, exposing the need for single diagnostic tests where TB testing is integrated with testing of other relevant and circulating respiratory illnesses capable of causing comparable clinical presentation and public detriment. New dual or multi-pathogen detection tests need to be accurate, rapid, affordable and accessible, particularly at primary healthcare centres with limited infrastructure and resources.

In this multi-centre diagnostic accuracy study in adults with signs and symptoms of TB, we determined that the multiplex Truenat MTB Ultima/COVID-19 assay was able to detect both MTB and SARS-CoV-2 in participants in a single assay. This study showed a sensitivity of 79.8% and specificity of 98.9% for detection of MTB when a sputum sample was mixed with a nasopharyngeal swab, and 64.4% sensitivity and 99.2% specificity for detection of COVID-19. Using an alternative sampling method of oral tongue swab mixed with a mid-turbinate nasal swab, sensitivity for both MTB and COVID-19 was lower, at 45.9% and 41.4% respectively, while maintaining high specificity. Although optimal diagnostic performance was not demonstrated, the potential and need for rapid development of tests that integrate TB diagnosis with the detection of other relevant respiratory infections is highlighted.

Traditional methods of TB diagnosis have limitations. Sputum smear microscopy has low sensitivity, typically detecting only 23–67% of pulmonary TB cases, particularly performing poorly in smear-negative and HIV-associated disease. TB culture, while more sensitive, requires specialised laboratory infrastructure and often takes several weeks to yield results, increasing the risk of diagnostic delay and patient loss to follow-up [12–14]. The shift in practice toward molecular tests for TB diagnosis and detection of rifampicin resistance has significantly advanced the pathway toward TB control by providing rapid and accurate diagnosis of both. However much more still needs to be done and even though current existing molecular tests, such as Xpert MTB/RIF and Ultra have demonstrated good performance, challenges such as the requirement for a temperature-controlled setting with a stable power supply have limited accessibility particularly in resource-constrained environments, where the burden of disease is highest [4,13]. The Truenat MTB, Truenat MTB Plus and Truenat MTB-RIF Dx chip-based PCR assays were developed to be run on the battery-operated, low-complexity, room-temperature-stable Truelab device as an alternative molecular assay to improve accessibility to TB diagnostics, particularly in resource-constrained settings where advanced diagnostic laboratories and skilled laboratory staff are scarce [12]. The Truenat MTB Ultima/COVID-19 chip was further developed to be run on the same Truelab device to facilitate both TB and COVID-19 detection using a single test.

This is not the first such study to show value of integrated TB and COVID-19 testing, but it is to our knowledge the first such study to show integrated testing for detection of both pathogens in a single multiplex assay. A prior study in Peru demonstrated the concept of concurrent TB and COVID-19 testing using a single sputum sample run across two separate assays on the same Xpert platform [15]. The authors showed the diagnostic yield of Xpert Xpress on sputum was moderate for COVID-19 (detected 67% of COVID-19 cases), and high for TB using Xpert Ultra (detected 96% of culture-confirmed TB cases). Despite this moderate performance using sputum for COVID-19, this study demonstrated the feasibility of combined and integrated testing.

In our study, the Truenat MTB Ultima/COVID-19 assay was developed to detect both MTB and COVID-19 with improved sensitivity in a single assay, using both sputum and swab samples together. Compared to Truenat MTB Plus, the Ultima chip newly includes both the *IS6110* and *IS1081* multi-copy gene targets to detect MTB, with a hypothetical improvement in sensitivity.

The overall performance of the Truenat MTB Ultima/COVID-19 assay using the combination of both sputum and a nasopharyngeal swab for the detection of MTB in this study was similar to prior reports for Truenat MTB Plus on sputum alone [16–18]. While one study showed increased sensitivity of the Truenat MTB Ultima assay for detection of TB, compared to MTB Plus, we failed to see the same enhanced performance here. The study by Abdulgader et al also showed a matched decrease in specificity, so more evidence may be required to evaluate if the new Ultima chip has improved performance overall [19].

However, when compared specifically for the detection of MTB in sputum alone on Xpert Ultra, the Truenat MTB Ultima/COVID-19 assay had 9.5% lower sensitivity. This may be related to innate assay design differences, operational parameters or population variance. Indeed, comparative performance varied across sites (Table 4), with the biggest difference seen in Peru and South Africa and the lowest in India and Uganda, suggesting difference by site may be due to variation in the patient populations sampled, quality of samples collected, or laboratory flow and processing of samples.

Several studies have shown marginally lower sensitivity of Truenat MTB Plus compared to Xpert Ultra when using sputum alone, although all are challenged by small sample sizes and restricted geographies [18,19]. This heterogeneity in TB diagnostic performance has also been seen with the Xpert assays [13]. In our study, the lower sample input volume was a likely contributing factor to lowered sensitivity: the total volume of sputum that was added to the Truenat MTB Ultima/COVID-19 assay was reduced, as there was a dilution effect when mixing 500 ul of sputum with the viral transport medium and the sample preparation buffer, and the assay itself is restricted to only a 6ul purified DNA input volume. We attempted to overcome this limitation by using an alternative sample collection approach, which we expected to be more acceptable to participants and without a large trade off in overall performance. By evaluating a combined oral tongue swab with a mid-turbinate nasal swab, where there was no concern about an inherent dilution factor, we saw reduced sensitivity of both MTB and COVID-19. This was not surprising for MTB, as there are published studies suggesting lower bacterial load on tongue swabs compared to sputum [20–22]. But it was surprising for COVID-19 where reports have shown largely comparable performance on midturbinate nasal swabs and nasopharyngeal swabs [23–25]. Nevertheless, the low sample size for clinically confirmed COVID-19 cases in this study precludes clear outcomes for detection of COVID-19.

Overall, the performance of the Truenat MTB Ultima/COVID-19 for COVID-19 detection in this study was unexpectedly poor. In previous diagnostic accuracy studies from India, performance as a single test was reported as having 100% sensitivity and 99-100% specificity [26,27]. In our own laboratory analytical study conducted in India, the performance of the Truenat MTB Ultima/COVID-19 chip for the detection of COVID-19 on biobanked combined nasopharyngeal swab and sputum was excellent (sensitivity 100%) against a nationally approved PCR test. We acknowledge that finding may be of limited utility given that this component of the study was only performed in India with a limited sample size (n=344) [28]. Nevertheless, it does suggest that the MTB Ultima/Covid-19 assay has excellent capacity to detect COVID-19, but that the sampling or population may have limitations.

The sensitivity of the Truenat MTB Ultima/COVID-19 for COVID-19 detection varied by site (48.3%-94.7%) and prevalence (0.7-5.9%) of COVID-19 (Table 4), with performance best in India, again highlighting, similar to TB diagnostic performance, the heterogeneity of results potentially being based on differences in populations, staff and site specific factors, as well as sample collection, laboratory flow and processing of samples. Other factors that may have also contributed to the assay’s poorer performance for COVID-19 detection in this study could be because the study was conducted later in the pandemic, with fewer people presenting for COVID-19 diagnosis (as demonstrated by the low overall COVID-19 prevalence), participants having lower viral load and shorter periods of viremia post COVID-19 vaccination [29], and because the study inclusion criteria were based on typical TB symptoms rather than typical COVID-19 symptoms.

While this assay is unlikely to have further clinical utility based on performance, it has demonstrated the potential of the rapid utilisation of newly developed multiplex diagnostic tools, particularly for respiratory infections that are a serious public health concern. Further learnings could be gained from optimising sample collection techniques and processing steps of mixed specimen samples.

## CONCLUSION

Although this study did not demonstrate optimal results for either TB (sensitivity was 79.8% and specificity was 98.9%) or COVID-19 (sensitivity 64.4%, specificity 99.2%) detection, it highlights the potential of rapid development and utilisation of tests that integrate TB diagnosis with diagnosis of other existing or new respiratory infections, mitigating the consequences of missed TB diagnosis particularly on a large scale like the COVID-19 pandemic did.

## Data sharing

The dataset is available from the corresponding author upon request. Upon acceptance, the dataset will be made available in the Dryad data repository, accessible at (to be updated before acceptance). The data has been shared under the public domain CC0 licence waiver.

## Data Availability

Data sharing: The full dataset is available from the corresponding author upon request. Upon acceptance, the dataset will be made available in the Dryad data repository, accessible at (to be updated before acceptance). The data has been shared under the public domain CC0 licence waiver.

## Acknowledgements

The authors would like to thank the study participants, their families and all staff at the study sites for their participation and contributions to the study, including those at CIDRI group based at Site B Khayelitsha, South Africa; health center in DIRIS Lima Centro and Hospital Huaycan in Lima, Peru; the Central Research Lab in Ujjain, India. The National Health Laboratory Services at Groote Schuur Hospital in South Africa provided space, resources, expertise and personnel to accommodate testing of specimens from the University of Cape Town. Acomed Statistik supported formal data analysis in this project. Mary Gaichiri and Alex Ogwal provided project management for this project. Sunita Singh supported clinical activities around the RD Gardi site. Michael Otieno supported development of the clinical database. We are grateful to Molbio Diagnostics for donation of the tests and reagents to conduct this study.

## Author contributions

AP-N, RS, and MR conceptualised the study. HT, RS, JS, WZ, MP and AP-N were responsible for data curation. Formal analysis was performed by HT, RS and AP-N. Funding acquisition was led by AP-N. Investigation was led by MJ, CU-G, MP, HC, CZ, WS. RS, MJ, MP, HC, CZ, WS and AP-N contributed to design of methodology. RS and AP-N were responsible for project administration. AP-N was responsible for the provision of resources. RS and AP-N supported software and data capture testing. RS, MJ, CU-G, MP, HC, CZ, WS and AP-N were responsible for supervision. Validation was performed by HT, RS and AP-N. HT, RS and AP-N were responsible for visualisation and data presentation. HT and AP-N lead writing of the original draft. All authors contributed to review and editing of the final manuscript.

## Generative AI statement

The author(s) declare that no Gen AI was used in the creation of this manuscript.

